# Using DeepLabCut for tracking body landmarks in videos of children with dyskinetic cerebral palsy: a working methodology

**DOI:** 10.1101/2022.03.30.22272088

**Authors:** Helga Haberfehlner, Shankara S. van de Ven, Sven van der Burg, Ignazio Aleo, Laura A. Bonouvrié, Jaap Harlaar, Annemieke I. Buizer, Marjolein M. van der Krogt

## Abstract

Markerless motion tracking is a promising technique to capture human movements and postures. It could be a clinically feasible tool to objectively assess movement disorders within severe dyskinetic cerebral palsy (CP). Here, we aim to evaluate tracking accuracy on clinically recorded video data.

**Method:** 94 video recordings of 33 participants (dyskinetic CP, 8-23 years; GMFCS IV-V, i.e. non-ambulatory) from a previous clinical trial were used. Twenty-second clips were cut during lying down as this is a postion for this group of children and young adults allows to freely move. Video image resolution was 0.4 cm per pixel. Tracking was performed in DeepLabCut. We evaluated a model that was pre-trained on a human healthy adult data set with an increasing number of manually labeled frames (0, 1, 2, 6, 10, 15 and 20 frames per video). To assess generalizability, we used 80% of videos for the model development and evaluated the generalizability of the model using the remaining 20%. For evaluation the mean absolute error (MAE) between DeepLabCut’s prediction of the position of body points and manual labels was calculated.

**Results:** Using just the pre-trained adult human model yielded a MAE of 121 pixels. An MAE of 4.5 pixels (about 1.5 cm) could be achieved by adding 15-20 manual labels. When applied to unseen video clips (i.e. generalization set), the MAE was 33 pixels with a dedicated model trained on 20 frames per videos.

**Conclusion:** Accuracy of tracking with a standard pre-trained model is insufficiently to automatically assess movement disorders in dyskinetic CP. However, manually adding labels improves the model performance substantially. In addition, the methodology proposed within our study is applicable to check the accuracy of DeepLabCut application within other clinical data set.

## Introduction

Movement disorders in dyskinetic cerebral palsy (CP), i.e. dystonia and choreoathetosis, are associated with impaired muscle tone regulation and interfere with intentional movements^1^. To treat movement disorders, neuromodulation treatments, including implanted medicine pumps and deep deep brain stimulation, aim to minimize the pathologic movements ^1^. Accurate and reliable measurements of the pathological movements are essential for indication and evaluation of these treatments. However, it remains a huge challenge to capture the complexity of the dyskinetic movement disorder in an objective way. Currently dystonia and choreoathetosis are assessed by a variety of clinical scales, most of which are video recorded and scored afterwards, such as the Dyskinesia Impairment Scale^2^ or Barry-Albright Dystonia Scale^3^. Major drawbacks of these scales are that they measure at one time point and mostly in a situation that is unfamiliar to the patient. In addition, outcomes of these scales are subjective, i.e. dependent on the personal judgement and experience of the rater. Therefore clinicians and researchers seek for other possibilities to objectively assess motor function in dyskinetic CP^4^. However, up to now there is no established instrumented clinical assessment method available for children and young adults within non-ambulatory individuals with dyskinetic CP.

We suggest a machine learning approach to automatically classify dyskinetic movement patterns, using data extracted by markerless motion tracking from 2D videos. In the long-term these videos can be recorded within the home-environment by caregivers. As a first step, here we aim to assess accuracy of tracking body landmarks from 2D videos compared to manual labeling in children and young adults with dyskinetic CP, using an open-source toolbox (DeepLabCut^5,6^) with a model pre-trained on a human healthy adult data set^7^ as backbone.

## Methods

### Participants

Ninetyfour videos of 33 unique participants of a trial on the effects of intrathecal baclofen (IDYS trial Dutch Trial Register NTR3642)^8^ from Amsterdam UMC, location VUmc, were used for the current analysis. Videos were collected at baseline and at two follow-up measurements within a year’s time. Patients at baseline had following characteristics: age 14.0 ± 3.9 (mean ± Standard deviation) years; weight: 32.7 ± 11.8 kg; height: 147.1 ± 20.4 cm; 8 females / 25 males; Gross Motor Function Classification System (GMFCS) IV (N=13) or V (N=20), Manual Ability Classification System (MACS): III (N=3), IV (N=8), V (N=22). This secondary analysis of the video data was approved by the local medical ethics committee.

### Video data

Sequences of twenty seconds (500 frames) were selected from videos in which children were lying in rest on a mat. This is a position that enables this group of non-ambulatory individuals to be assessed without external support. Within all videos the faces of the children and caregivers were blurred using Mondrian (version 1.1, Kinedata), a custom-made semi-automatic program. The face blurring was necessary to fulfill the privacy regulations of the Amsterdam UMC for the succeeding analysis using DeepLabCut within a cloud environment provided by SURFcumulus (SURF, Utrecht, the Netherlands) based on an Azure cloud environment (Microsoft Azure, Europe West). Subsequently, the videos were all converted to the same size (720 width x 575 height pixels, which covered an area of about 3×2 meter, i.e. an image resolution of 0.4 cm per pixel), and the same video format (avi, svd2500 codec) using Any Video Converter (version 5.7.8, Anvsoft Inc).

We split the data in a development set and a generalization set. 80% of the participants (i.e. 27 subjects with 76 related videos) were randomly placed in the development set and 20% of participants (i.e. six participants with 18 related videos) in the generalization set. The development set was used to train models with an increasing number of manually labeled frames (as explained in detail below). The videos of the generalization set were manually labeled as well, but kept apart from the model development process to show the potential of generalizability towards “unseen” videos. The process of splitting the data and the following processing in DeepLabCut is visualized in Figure 1.

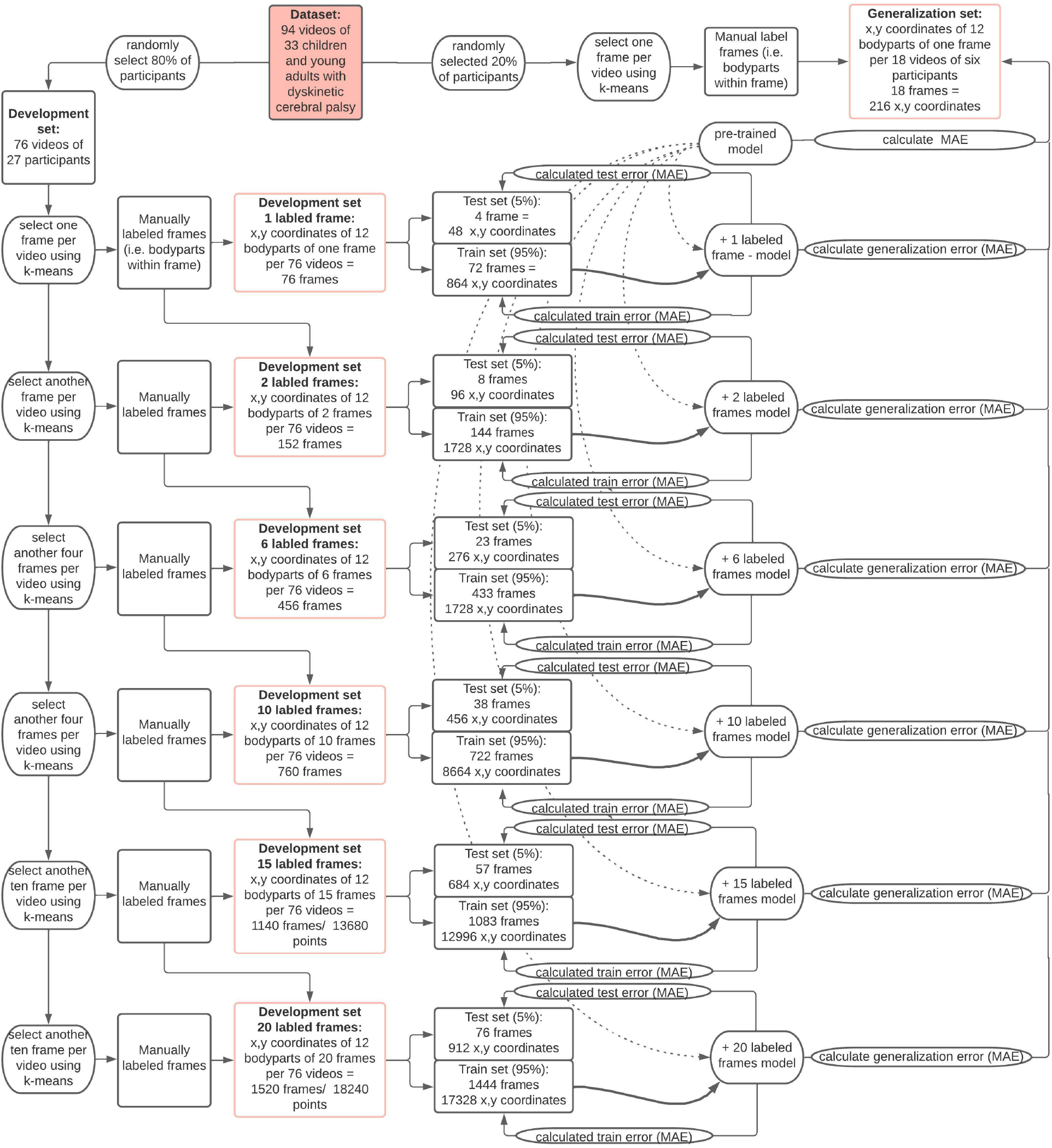

### DeepLabCut

DeepLabCut is a recently developed open-source toolbox that allows training of a deep neural network using pre-trained models with limited training data to track user-defined body points using transfer learning^5,6^. We ran DeepLabCut (Version 2.1) using a single NVIDA Tesla K80 GPU platform via Microsoft Azure’s cloud with a ‘Data science Virtual Machine - Windows 2019)’ blueprint. The conda environment (for GPU provided by DeepLabCut) was used within a Jupyter notebook. Models were trained using an available residual neural network with 101 layers (ResNet-101) pre-trained on the MPII Human pose dataset^7,9,10^ as initial weights. This human model is available as built-in option in DeepLabCut. The same body points included in MPII (i.e. wrists, elbows, shoulders, hips, knees and, ankles) were manually labeled within the development set up to 20 frames per videos with exception of chin and forehead (these body points could not be labeled due to blurring of the faces). One frame per video was labeled within the generalization set. Frames for labeling were automatically selected beforehand by DeepLabCut using k-means clustering to select frames with a variety of postures within the datasets. The labeled frames of the development set were randomly split in a training and test set (95% training dataset, 5% test dataset). The amount of training and test data is presented in Figure 1 for each dataset. Different models were first trained on the development set with an increasing number of manually labeled frames (1, 2, 6, 10, 15 and 20) (Figure 1). Training was performed using the default settings of DeepLabcut e.g. shuffle is true. All models were trained up to 400.000 iterations with a batch size of one. The human model was always taken as the intial weight within the training. The graphs of cross-entropy loss were inspected to determine convergence and define minimal training iterations needed for our dataset.

### Evaluation

We evaluated all models towards their own dataset, i.e. test and train error, and towards the generalization data set, i.e. generalization error (Figure 1). The model evaluation was performed within DeepLabCut by calculation of the Euclidean distance for the pairs of x,y coordinates (i.e. manually labeled versus predicted by the model). The mean of Euclidean distances (across all body points and frames) was taken as the mean absolute error (MAE). MAEs were calculated with and without a p-cutoff of 0.8 (i.e. leaving predicitions out with a low likelihood to be correctly identified by the model).

## Results

Within the test set MAE decreased from 10.09 (1 labeled frame) to 4.49 pixels (20 labeled frames) (Table 1). Applying a p-cutoff of 0.8 did not significantly affect the error. The lowest MAEs were reached with the model with 15-20 additional labeled frames per video. Within the generalization set MAE decreased from 107.04 (no labeled frame – i.e. pre-trained model only) to 33.18 pixels (20 labeled frames) (Table 1). Applying a p-cutoff within the genberalization set improved the MAE towards 19.88 pixels (Table 1).

**Table 1.:** Mean averaged error (MAE) in development set (train, test) and in the generalization set, evaluated with and without p-cutoff 0.8

| Labeled frames | Train set | Test set | Test set with p-cutoff 0.8 | Generalization set | Generalization set with p-cutoff 0.8 |
| --- | --- | --- | --- | --- | --- |
| 0 |  |  |  | 107.04 pixels | 121.18 pixels |
| 1 | 1.17 pixels | 10.09 pixels | 10.23 pixels | 37.24 pixels | 28.72 pixels |
| 2 | 1.11 pixels | 5.83 pixels | 5.83 pixels | 39.00 pixels | 28.89 pixels |
| 6 | 1.65 pixels | 5.14 pixels | 5.06 pixels | 32.82 pixels | 23.51 pixels |
| 10 | 1.8 pixels | 5.86 pixels | 5.18 pixels | 34.68 pixels | 25.91 pixels |
| 15 | 2.36 pixels | 4.49 pixels | 4.48 pixels | 36.26 pixels | 28.33 pixels |
| 20 | 2.71 pixels | 4.49 pixels | 4.48 pixels | 33.18 pixels | 19.88 pixels |

## Discussion

DeepLabCut provides an accessible platform for tuning models towards an own video dataset, which is necessary for pathologic movements not included in standard human datasets, especially in postions not commonly include in pretrained datasets such as lying down. Within this work we show how to evaluate the accuracy of DeepLabCut application on a clinical data set. We assume that (1) spliting an available dataset within a development set and generalization set, (2) applying an increasing number of manual labels and (3) independenly assess the tracking error (MAE) is a suitable approach. The same approach (Figure 1) can also be used towards new, different datasets.

We achieved a tracking error of 1.5 cm, which did not further improve between 15-20 labeled frames per video. Within the assessment of dystonia and choreoathetosis we suggest that general movement and posture features will be required, such as movement frequency and distance of bodypoint towards the body center. Although the accuracy may effect this features, we assume that the accuracy is sufficient for our planned approach to use this body landmarks for the assessment of dystonia and choreoathetosis.

We did not systematically compare DeepLabCut with other options for human pose estimation such as OpenPose^11^ or the most recently developed EfficientPose^12^. Within a pilotphase of our project, we also tried OpenPose but the results (assessed by visual inspection of the overlayed videos) did not show sufficient tracking results for automatic tracking within our dataset. Anyhow, for all tracking options we deem it necessary to evaluate accuracy of tracking when used in pathologies in childhood as models have mainly developed withing abled bodied adults without taking into account bony deformities or environmental factors such as a wheelchair or walker in the picture. In addition, “crowdsource labeling” as used for MPII dataset, might not fulfill the requirements of anatomical representation of the body part, as recently highlighted in a review of Cronin et. al (2021)^13^. Therefore, transfer learning using manual labeling can be also useful in situations where exact anatomical positions are needed for further processing.

One major problem in the movement analysis in dyskinetic CP is small sample size. By using markerless motion tracking from 2D videos, historical data could be used for model development and extracted body position could be easily pooled without privacy issues. In this way, markerless motion tracking might open possibilities towards development of conventional machine learning or deep learning approaches for the assessment of complex movement disorders such as dyskinetic CP. Similar methodologies are currently developed for generalized movement analysis^14^ and automatic gait assessment in CP^15^.

## Conclusion

Markerless motion tracking using DeepLabCut with a standard pre-trained model is insufficiently accurate to automatically track body points in children and young adults with dyskinetic CP. However, accuracy substantially improves by adding manual labels towards 15-20 frames per video. This opens up possibilities for near-automatic tracking of videos. The proposed methodology is applicable to assess and improve this tracking accuracy for different clinical populations.

## Data Availability

Raw data of this study are patient videos and can not be made available. The outcome data are available online as data_stickfigure_coordinates_lying.zip within the MODYS-video dataset at zenodo https://doi.org/10.5281/zenodo.5638470

https://doi.org/10.5281/zenodo.5638470

## Funding

The project is funded by the Netherlands Organization for Health Research and Development (ZonMW, Innovative medical device initiative (IMDI) project number 104022005) Helga Haberfehlner is funded by the Postdoctoral Fellow Marie Skłodowska-Curie Actions - Seal of Excellence of the Research Foundation – Flanders (SoE fellowship_12ZZW22N).

## Notes

### Competing Interest Statement

The authors have declared no competing interest.

### Author Declarations

This secondary analysis of the video data was approved by the local medical ethics committee (METC VU medical center)

